# Breaching the curation bottleneck with human-machine reading symbiosis

**DOI:** 10.1101/2021.07.14.21260440

**Authors:** Cliff Wong, Rajesh Rao, Taofei Yin, Cara Statz, Susan Mockus, Sara Patterson, Hoifung Poon

## Abstract

**Purpose:** The explosion of molecular biomarker and treatment information in the precision medicine era drastically exacerbated difficulty in identifying patient-relevant knowledge for clinical researchers and practitioners. Curated knowledgebases, such as the JAX Clinical Knowledgebase (CKB) are tools to organize and display knowledge in a readily accessible format; however, curators face the same challenges in comprehensively identifying clinically relevant information for curation. Natural language processing (NLP) has emerged as a promising direction for accelerating manual curation, but prior applications were often conceived as stand-alone efforts to automate curation, and the scope is often limited to simple entity and relation extraction. In this paper, we study the alternative paradigm of assisted curation and identify key desiderata to scale up knowledge curation with human-computer symbiosis.

**Methods:** We chose precision oncology for a case study and introduced self-supervised machine reading, which can automatically generate noisy training examples from unlabeled text. We developed a curation user interface (UI) for precision oncology and through iterative “curathons” (curation hackathons), conducted retrospective and prospective user studies for head-to-head comparison between manual and machine-assisted curation.

**Results:** Contrary to the prevailing assumption, we showed that high recall is more important for end-to-end assisted curation. In extensive user studies, we showed that assisted curation can double the curation speed and increase the number of findings by an order of magnitude for previously scarcely curated drugs.

**Conclusion:** We demonstrated that an iterative and thoughtful collaboration between professional curators and NLP researchers can facilitate rapid advances in assisted curation for precision medicine. Human-machine reading symbiosis can potentially be applicable to clinical care and research scenarios where curation is a major bottleneck.

## Introduction

A perennial challenge to clinical researchers and practitioners is how to stay abreast with the rapid growth of biomedical research literature. This problem is particularly acute in the era of precision medicine, with an explosion of knowledge about the myriad molecular biomarkers and treatments. Manual curation requires domain expertise and is expensive and time-consuming. Natural language processing (NLP) has emerged as a promising direction for accelerating curation. However, prior work tends to consider NLP as a stand-alone approach for automatic curation. The scope is often limited to entity search or simple relation extraction (e.g., binary relations within single sentences), focusing on extracting common findings with high precision^1,2^.

In this paper, we study the alternative paradigm of assisted curation, where curation is approached as an interactive collaboration between human experts and NLP-based machine readers. Machine readers can scan millions of documents in seconds, reducing the giant haystack to a small set of candidate needles. Such candidate facts are presented in a curation user interface (UI), which human experts can quickly verify by essentially clicking a button. The curation decisions provide feedback to further improve machine reading accuracy.

The emphasis on human-computer symbiosis has prompted several significant design changes. Attaining high coverage (i.e., high sensitivity/recall) is of paramount importance in machine reading, as we are striving to leave no facts behind. Precision (i.e., positive predictive value) still matters for curation efficiency, but even modest precision such as 50% can lead to substantial efficiency gain, while ensuring excellent coverage in the long tail of scientific findings. This opens up opportunities to leverage the latest progress in NLP and machine learning, such as self-supervised machine reading, which uses prior knowledge to automatically generate noisy training examples from unlabeled text.

We conduct a case study on precision oncology and show how an iterative and thoughtful collaboration by professional curators and NLP researchers has led to rapid advances in assisted curation for molecular tumor boards and precision medicine. Motivated by end-to-end curation needs, we expanded the scope of self-supervised machine reading to cross-sentence, document-level extraction of complex n-ary relations, compared to prior work that often focuses on extracting entities or simple relations^3^. Without any annotated examples, our self-supervised machine reader already attains high recall and reasonable precision. We developed a curation UI for precision oncology through iterative “curathons” (curation hackathons). In extensive user studies, we show that assisted curation can double the curation speed in general and increase the number of findings by an order of magnitude for previously scarcely curated drugs.

## Methods

Precision oncology has seen an explosion of new findings on how tumors with a certain genomic biomarker (e.g., a point mutation or fusion) might respond to a given treatment. Manual curation of such knowledge typically starts with an explicit triaging step, e.g., by querying the PubMed search engine with relevant keywords such as “cancer”, “mutation” and a drug name. Each query might return hundreds of PubMed papers for a given drug and curation of each paper can take hours.

In assisted curation, we instead apply a machine reader to scan all PubMed articles and identify candidate findings, which curators can verify in a user interface (Figure 2). Each relation candidate is shown in context, with relevant entities highlighted. Users can also check the full article for more information. Curators can simply click a button to accept or reject the proposed extraction, thus greatly accelerating curation. The machine reading system and the assisted curation UI both stem from Project Hanover in Microsoft Biomedical NLP, and we refer to them as Hanover in this paper.

**Figure 1:**
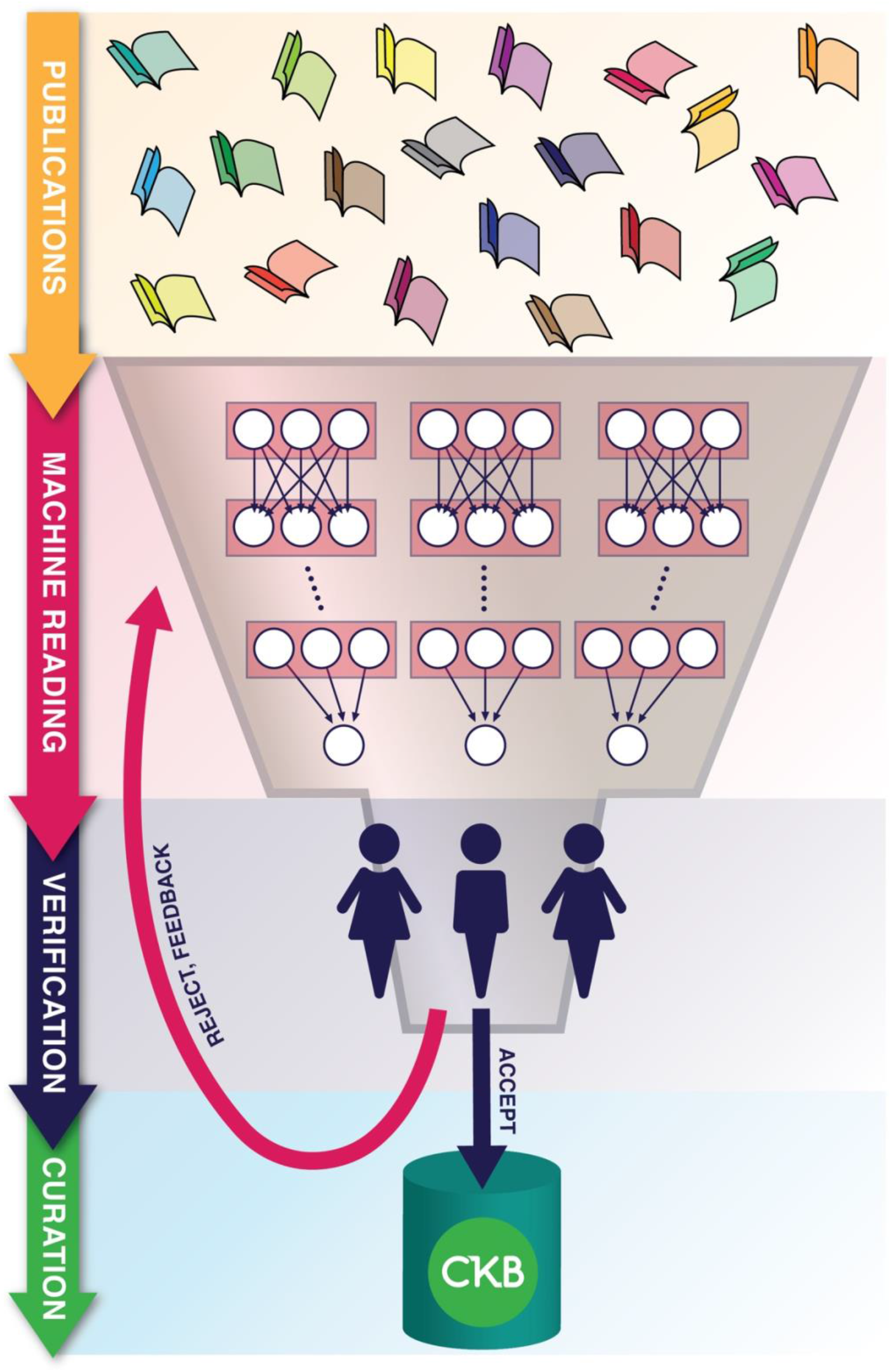
Assisted curation applies machine reading to reduce a giant haystack of source documents to a few candidate findings, which human experts can quickly validate through a curation user interface.

**Figure 2:**
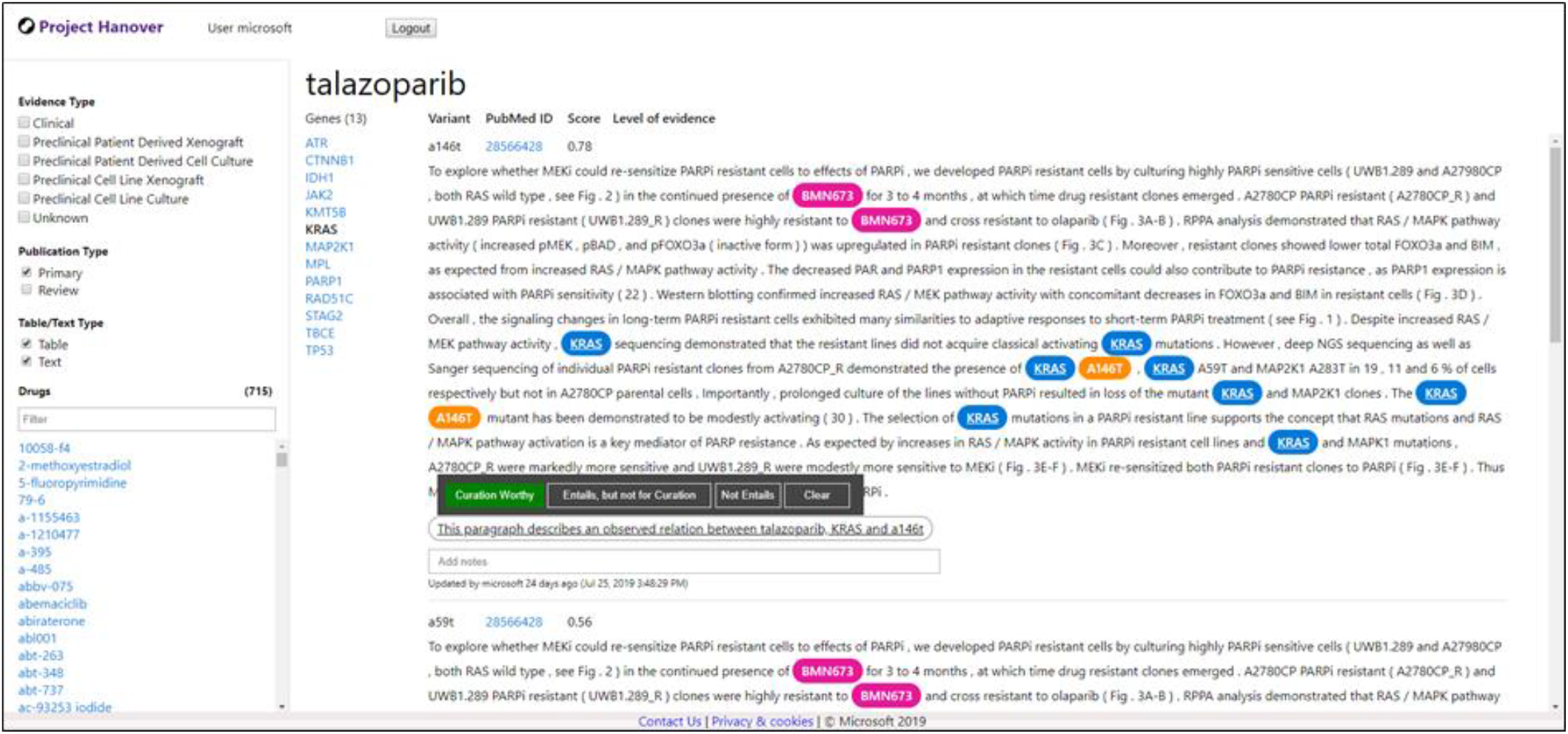
Assisted curation user interface for precision oncology. Users can verify a candidate finding (how a tumor with a given gene mutation might respond to a drug) by clicking a button.

### Self-supervised machine reading

The machine reader is powered by an NLP system that learns to extract such findings from text. Standard information extraction focuses on entity search or binary relations within single sentences. In precision oncology, as in many other biomedical scenarios, we need to extract complex n-ary relations that may span multiple sentences, such as the aforementioned drug-gene-mutation relation, which can be framed as a classification problem (whether a given text span mentions drug response given tumor gene mutation for a drug-gene-mutation tuple). Standard supervised learning requires manually annotating a large number of input-output examples. Instead, we apply self-supervised machine reading, which leverages an available knowledge base (KB) of known drug-gene-mutation relations to automatically annotate noisy training examples in unlabeled text. For any high-value relation, manually curated KBs generally exist. While manual curation is hard to sustain and can produce sparse coverage, these KBs can be used as a seed to bootstrap machine reading. Such a self-supervised approach has proven effective in our prior work^4^, and we use the same method in this paper. Specifically, we use three public KBs (CIVIC^5^, OncoKB^6^, GDKD^7^) for self-supervision, which contain 1,256 unique drug-gene-mutation relation tuples. For unlabeled text, we use the PubMed Central Open Access Dataset, accessed in June 2018, with 1.65 million full-text articles. To identify drug and gene mentions, we use a curated list of canonical names and aliases based on publicly available ontologies. For mutations, we use simple regular expressions, focusing on missense mutations in this study. For training, we identify drug-gene-mutation tuples that appear nearby in text and automatically label an instance as positive if the tuple is in the KBs and negative otherwise. We focus on co-occurring mentions within a paragraph and leave cross-paragraph extraction to future work. We subsample the negative instances to ensure a balanced training set. This yields 4,906 positive and 13,371 negative examples, respectively, which we use to train a recurrent neural network (long short-term memory or LSTM).

To support end scenarios such as molecular tumor boards, curators might need more than drug-gene-mutation relations, such as the evidence type and strength (e.g., whether the finding stems from a human trial, the sample size, etc.). An automated curation system would have to tackle all these required aspects at once. In assisted curation, however, it suffices to focus on the core drug-gene-mutation relation initially, as curators can easily check the additional aspects during verification. This can already attain the majority of efficiency gain in curation, as most irrelevant papers do not contain any mention of the core relation.

### User study

We applied the self-supervised machine reader to PubMed-scale knowledge extraction and assessed extraction quality and potential efficiency gain from assisted curation. For accuracy estimate, we sampled the machine-reading results and conducted several curathons with professional curators of the JAX Clinical Knowledgebase (CKB)^8^ to annotate whether a candidate drug-gene-mutation relation instance was “Curation Worthy”, “Entails, but not for curation”, or “Not Entail”. A candidate relation is considered “Curation Worthy” when it is described in the text (i.e., machine reading is correct), and it is deemed valid for curation into the database by the curators. “Entails, but not for curation” indicates that while the candidate relation is indeed described in the text, it is deemed not fit for curation given additional context in the paper (e.g., insufficient evidence strength). “Not Entails” indicates the candidate relation is not described in the text (i.e., machine reading is wrong).

For coverage estimate, we identified unique drug-gene-mutation ternary relations in machine reading results and compared them with those in CKB. For relations not in CKB, we used the above sample precision to estimate the curation-worthy portion. We conducted a series of controlled user studies to estimate potential efficiency gain from assisted curation. To ensure fair comparison, two curators with comparable curation efficiency and familiarity with both curation methods participated in the study. Both manual curation and assisted curation focus on missense mutations and PubMed and PMC text published in or after 2014.

## Results

In each curathon and user study, we exclude relations that are already curated in CKB. In total, CKB curators have annotated 1147 candidate drug-gene-mutation relation instances from self-supervised machine reading (Hanover). 749 are considered entailed, among which 351 are deemed curation-worthy. Consequently, we estimate that machine reading attains a precision of 65.3% and a curation-worthy proportion of 30.6%. Generally, CKB curators only need 1-2 minutes to validate each instance using the assisted curation UI (Figure 2).

Overall, we have applied machine reading to all publicly available PubMed and PMC papers, including 32.4 million abstracts and 3,671,360 full-text articles. This yields 27,308 unique ternary drug-gene-mutation relations, with 2,878 already appearing in CKB and 7,329 among the remainder estimated to be curation-worthy. Figure 3 compares curation-worthy coverage by CKB manual curation and Hanover machine reading from publicly available PubMed papers over the years. In total, Hanover extracts an estimated 10,207 unique curation-worthy relations, compared to 4,493 in CKB curated from the same set of PubMed text, with an estimated addition of 7,329 new curation-worthy relations, or 163% increase in coverage.

**Figure 3:**
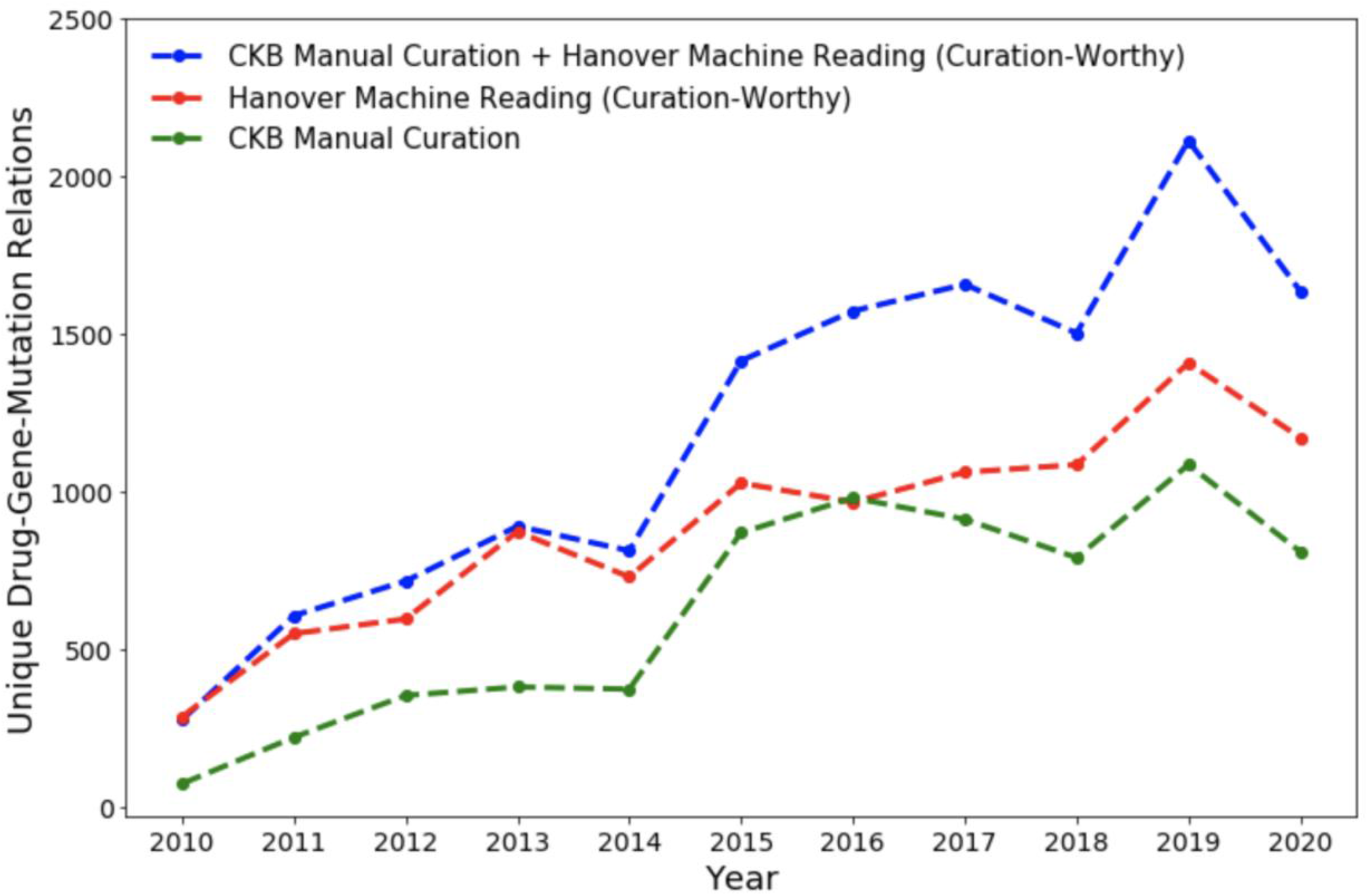
Comparison of coverage by Clinical Knowledgebase (CKB) and self-supervised machine reading (Hanover), as measured by unique drug-gene-mutation relations curated from PubMed articles over the years. If a finding appears in multiple publications, the first will be counted. The apparent dip in 2020 is due to embargo (delay in PMC release of full-text articles).

**Figure 4:**
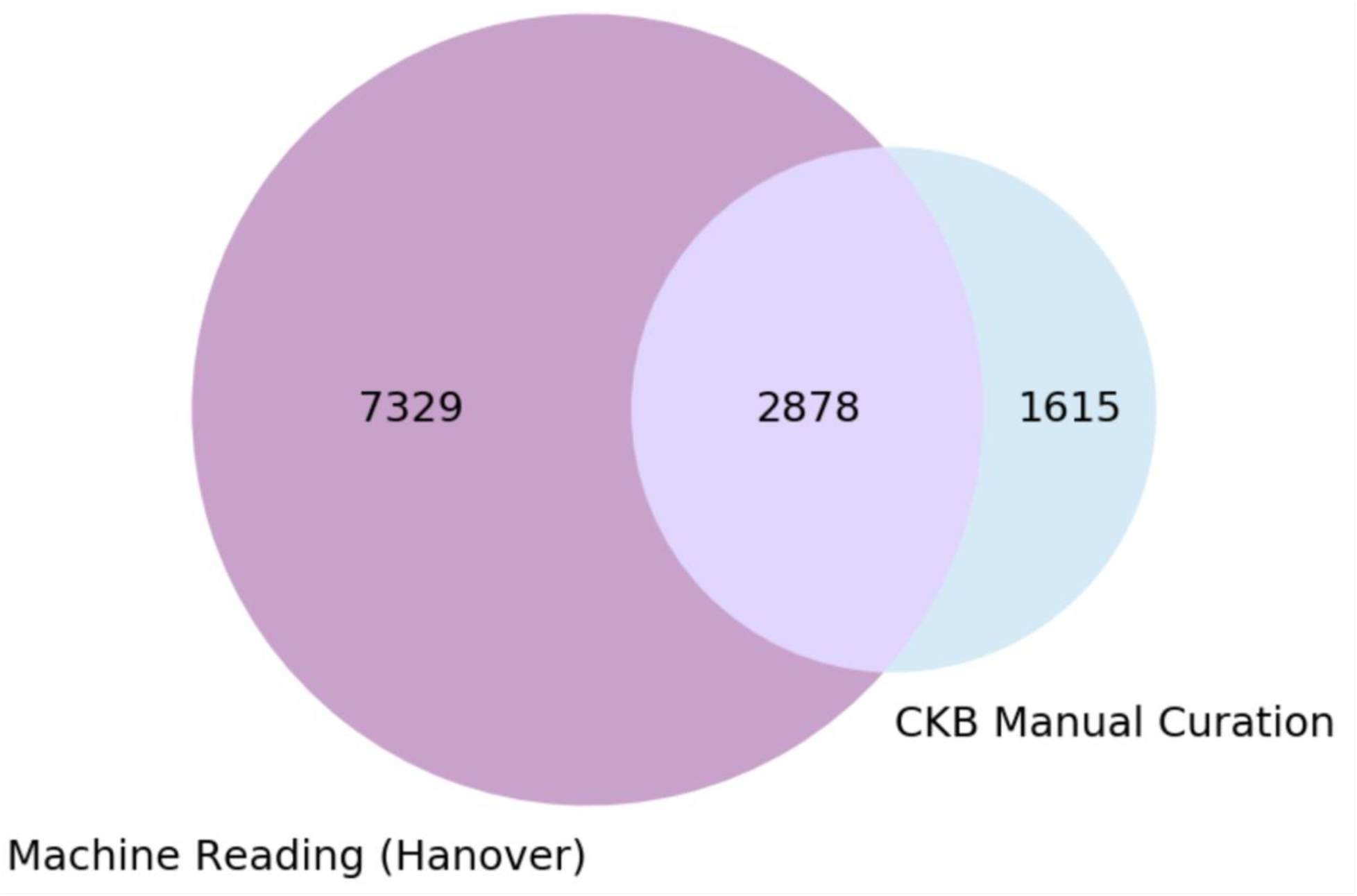
Comparison of curation coverage from publicly available PubMed articles, in unique drug-gene-mutation relations (machine reading with estimated curation-worthy results).

Finally, we conduct a series of controlled user studies to compare the efficiency of manual and assisted curation. We select 27 drugs that are relatively new and less well curated. In each user study, we always allotted equal time for manual and assisted curation. In the first two studies, the curators can choose the drugs to work on freely. In the final user study, we also pre-specify the allotted time for each drug in both manual and assisted curation, to enable more head-to-head comparison. The allotted time is set based on estimated abundance of findings. We set a minimum of 30 minutes for each drug, as manual curation requires significant time to conduct the initial triaging step. Assisted curation does not have such overhead and for a few drugs curators finished validating all candidate instances ahead of the allotted time. Table 1 shows the detailed results. Assisted curation doubles the speed, yielding a total of 351 unique findings, compared to 180 in manual curation. It also uses less time, 669 vs 700 minutes. As expected, manual curation covers far fewer PubMed articles compared to assisted curation, 78 compared to 221 articles. While assisted curation already doubles the speed, there is much room for further improvement. Often, manual curation benefits from curating a large number of relations from a single paper (e.g., 14 ternary relations from Table 9 in PMID 22734072 for drug tretinoin). Our current assisted curation UI, however, organizes candidate relations in the order of drug, gene, mutation. Therefore, findings from one paper might be scattered around based on the associated drugs and genes. A paper-centric view might enable much faster assisted curation.

**Table 1:** Comparison of manual and assisted curation efficiency in controlled user studies. In the first two studies, curators were given equal time (8 hours) for manual and assisted curation, respectively. In the final user study, we also pre-specified the allotted time for each drug to make the comparison more head-to-head.

| **Drugs** | **Allotted Time (min)** | **Manual Yield** | **Assisted Curation Yield** | **Machine Reading Precision (%)** | **Machine Reading Curation-Worthy Proportion (%)** |
| --- | --- | --- | --- | --- | --- |
| **User Study 1** |  |  |  |  |  |
| anastrozole | - | 7 | 7 | 75.0 | 25.0 |
| axitinib | - | 18 | 27 | 84.6 | 51.9 |
| bafetinib | - | 1 | 21 | 97.4 | 55.3 |
| bevacizumab | - | 8 | 2 | 27.6 | 6.9 |
| alectinib | - | 2 | 12 | 50.0 | 33.3 |
| alvespimycin | - | 0 | 33 | 70.7 | 56.9 |
| bortezomib | - | 0 | 32 | 56.9 | 31.4 |
| exemestane | - | 1 | - | - | - |
| bosutinib | - | 7 | - | - | - |
| **User Study 2** |  |  |  |  |  |
| tretinoin | - | 38 | 28 | 61.6 | 32.6 |
| vorinostat | - | 5 | 37 | 63.0 | 37.0 |
| tanespimycin | - | 10 | 21 | 81.0 | 33.3 |
| trastuzumab<br>emtansine | - | 4 | 0 | 60.0 | 0.0 |
| rociletinib | - | 2 | 3 | 94.1 | 17.7 |
| valproic acid | - | 3 | 6 | 65.3 | 12.2 |
| Sp600125 | - | 4 | - | - | - |
| **User Study 3** |  |  |  |  |  |
| sp600125 | 90 | 3 | 1 | 13.9 | 2.8 |
| infigratinib | 30 | 2 | 7 | 80.0 | 46.7 |
| exemestane | 30 | 3 | 8 | 100.0 | 21.6 |
| rituximab | 45 | 0 | 10 | 56.7 | 33.3 |
| epigallocatechin gallate | 45 | 2 | 5 | 79.1 | 11.6 |
| bosutinib | 60 | 2 | 4 | 63.5 | 7.7 |
| pazopanib | 60 | 1 | 18 | 56.3 | 37.5 |
| erlotinib | 90 | 11 | 4 | 48.0 | 5.3 |
| crenolanib | 30 | 8 | 16 | 86.4 | 72.7 |
| bortezomib | 60 | 8 | 8 | 51.4 | 21.6 |
| icotinib | 30 | 6 | 6 | 85.0 | 30.0 |
| quizartinib | 45 | 17 | 9 | 52.4 | 42.9 |
| genistein | 30 | 1 | 0 | 71.4 | 0.0 |
| navitoclax | 45 | 3 | 17 | 80.0 | 68.0 |
| tanespimycin | 30 | 3 | 9 | 82.4 | 52.9 |
|  |  | **180** | **351** |  |  |

Moreover, as mentioned before, while our machine reading precision is quite high (65.3%), the curation-worthy proportion is substantially lower (30.6%). This is because the current scope focuses on the core drug-gene-mutation relation, which does not account for additional aspects such as evidence type and strength. This suggests additional opportunities to improve assisted curation efficiency by expanding the scope of machine reading.

## Discussion

Our study demonstrates the promise of machine-reading assisted curation in accelerating knowledge curation as in precision oncology for molecular tumor boards. Machine reading substantially increases curation coverage, and curators no longer need to undergo a repetitive and time-consuming process to scan a large swath of mostly irrelevant text. Instead, they can start with candidate relations with provenance and quickly validate them in a curation-friendly UI, which can already double the curation speed. There are many opportunities to further increase the efficiency gain. Prior work in biomedical knowledge extraction, including commercially available tools, often focuses on entity search or simple relations such as binary drug-gene relations. For complex knowledge curation, as in precision oncology, they are typically insufficient to provide a significant boost in curation efficiency. For example, for one example drug, an entity search approach will yield over 800 PubMed articles that mention the given drug, as well as some gene mutations in the cancer context. Curating them all would have required over a thousand expert hours. By contrast, our machine reading system identifies 38 candidates, which curators finish validating in about an hour and identify 21 curation-worthy findings, whereas previously CKB only contained two findings for the given drug.

There are several limitations in our study. First, our text corpus is limited to publicly available PubMed text. There is much more biomedical text behind the paywall and applying machine-reading assisted curation to them can potentially extract even more knowledge. For simplicity, we focus on point mutations in this study, but in preliminary results we find that machine reading is also effective in extracting complex variants, such as fusion and indel. We focus on the core drug-gene-mutation relations in the initial exploration, but machine reading can be further expanded to include other aspects that are important for ascertaining curation-worthiness.

## Data Availability

Full-text publication data used for machine reading was from the PubMed Central Open Access Dataset (PMC) https://www.ncbi.nlm.nih.gov/pmc/. The full dataset for the results can be made available upon request.

